# Assessing sustainability factors for rural sanitation coverage in Kenya, Zambia, Nepal, and Bhutan: A qualitative case study analysis

**DOI:** 10.1101/2021.08.24.21262558

**Authors:** Zoe Sakas, Eberechukwu A. Uwah, Jedidiah S. Snyder, Joshua V. Garn, Matthew C. Freeman

## Abstract

**BACKGROUND:** Few countries are likely to achieve universal sanitation within the next decade as sustaining sanitation coverage remains a critical challenge. The purpose of this study is to investigate factors that may have supported or hindered sustainability of sanitation coverage 1-2 years after the completion of an integrated, area-wide sanitation program in four countries.

**METHODS:** Between 2014 and 2018, the SSH4A approach was implemented in 15 countries in Africa and Asia, four of which are included in this qualitative study. We conducted focus group discussions and interviews with beneficiaries, implementors, and decision-makers to identify sustainability factors and used household survey data to characterize sub-national sanitation coverage throughout implementation, and 1-2 years after.

**RESULTS:** Our data revealed behavioral, contextual, and service delivery factors that were related to the sustainability of sanitation improvements. Service delivery factors included follow-up hygiene promotion, access to materials (e.g., plastic, cement), local government commitment post-implementation, functioning monitoring systems, uptake of the supply chain by private sector, capacity for innovation. Contextual and behavioral factors included poverty, soil type, road networks, social cohesion, desire for improved latrines, maintenance and cleaning, and knowledge of sanitation benefits.

**DISCUSSION:** The presence or absence of sustainability factors identified through this research may have implications on where certain programmatic approaches will work, and where adaptations may be required. Through comparing sustainability factors with sub-national slippage rates, we were able to illustrate how local service delivery systems may respond to barriers (e.g., poverty, lack of affordable sanitation options, changes in population density) and enablers (e.g., sufficient resource allocation, passionate leadership, social cohesion). Understanding the programmatic and contextual factors that either drive or hinder long-term sanitation coverage may allow for greater program impact through adapting implementation based on existing challenges in service delivery and context.

## 1. Background

Achieving universal access to basic sanitation and hygiene remains challenging in low and middle-income countries (LMIC).^1–3^ Globally, 1.7 billion people lack basic sanitation facilities and few countries are projected to accomplish universal coverage within the next decade, especially as sustainability remains a key challenge for sanitation and hygiene interventions.^4–8^ Following these concerns, understanding sustainability factors for sanitation coverage has become a critical priority within the water, sanitation and hygiene (WASH) sector.^9–10^

To address the challenge of sustaining sanitation coverage in LMICs, interventions often aim to strengthen existing local service delivery systems through capacity building and technical assistance.^11–13^ International organizations are also starting to overtly address weaknesses within existing service delivery systems through engaging with local stakeholders including government officials, community leaders, and representatives in the private sector.^14^ Moreover, continuous political will, resource allocation, and prioritization of WASH within local governments ensures that improvements in sanitation service delivery systems are sustained post-intervention.^14, 15^

Understanding the programmatic and contextual factors that either drive or hinder long-term sanitation coverage may allow for greater program impact through adapting implementation based on existing challenges in service delivery and context. According to previous studies, local ownership and responsibility for sanitation services tends to support sustainable coverage, including government commitment and local capacity (e.g., resources, training) to innovate and adapt service delivery. Additionally, contextual and behavioral factors often lead to adaptations or improvements in service delivery based on the unique needs or challenges within a particular setting.^16–19^ In a recent report by USAID, ten contextual factors were quantitatively identified as having an impact on the sustainability of sanitation achievements, including: village size, access to improved water, population density, shrubland coverage, remoteness, forest coverage, literacy, distance to water bodies, waterborne disease burden and water scarcity.^20^ International organizations may assess existing service delivery and contextual factors through formative research in order to inform the design and implementation of sustainable sanitation interventions.

The Sustainable Sanitation and Hygiene for All (SSH4A) approach was implemented by SNV, an international development agency based in the Netherlands, in 15 countries between 2014 and 2018.^21^ The SSH4A approach focuses on four key components of rural sanitation programming: 1) demand creation, 2) supply chains and financing, 3) behavior change communication, and 4) WASH governance, and was designed within the context of local government planning and budgeting to promote sustainability post-intervention.^8, 22^ A recent assessment of the sustainability of sanitation improvements from SSH4A programming in ten countries indicated that there was sustained coverage in half of the program areas (Bhutan, Ghana, Kenya, Nepal, Tanzania) and varied levels of slippage in the others (Ethiopia, Indonesia, Mozambique, Uganda, Zambia) ranging from a drop of 63% in coverage in one program area in Ethiopia to a drop of only 4% in Indonesia.^23^ According to this study, factors associated with sustainability through quantitative analysis of household survey data included: household socioeconomic status, baseline sanitation coverage before SNV started working in the area, and the rate of change of sanitation coverage during program implementation period.^23^

The purpose of this study is to further explore factors that may have supported or hindered sustainability of national and sub-national coverage 1-2 years after implementation. We also examined the relationship between identified sustainability factors and varying levels of slippage in SSH4A program areas using qualitative key informant interviews (KII) and focus group discussions (FGD) with in-country project stakeholders, implementers, and beneficiaries.

## 2. Methods

We conducted qualitative case study analysis to identify factors related to the sustainability of sanitation coverage in four case studies (Kenya, Zambia, Nepal, and Bhutan) 2 years after completion of SSH4A activities in select program areas.

### 2.1. Country and district selection for case studies

This study was nested within an evaluation of a 5-year multi-country sanitation program. As part of this evaluation, repeated cross-sectional household surveys were administered throughout implementation and 1-2 years after the intervention was completed, to assess the sustainability of sanitation coverage gains. Details of the purpose and methods for this evaluation and the quantitative analyses are published elsewhere.^23^ Briefly, a multi-stage cluster sampling scheme was used to select a random and representative sample of 12 program areas representing 10 countries. Data were collected on household WASH access and use, including direct observations of sanitation facilities. For this study, we aggregated the household data at the district level, to show district-level sanitation trends over time. Our study team performed analyses to assess impact and sustainability of key household WASH variables, and these quantitative findings were used both to select study sites and to inform our qualitative findings.^8, 23^

We selected program areas from 4 of the 10 countries for further qualitative research. Countries were chosen based on having diversity in sanitation facility coverage improvements and post-intervention slippage, and qualitative factors including diversity, implementation variety, toilet quality, national policies, and geography.^8, 23^ Table 1 outlines geographic factors, population density, and livelihood in each of the included areas.

**Table 1.** Observed contextual factors from four case studies.

| Country | Kenya |  | Zambia |  | Nepal | Bhutan |  |
| --- | --- | --- | --- | --- | --- | --- | --- |
| Region | Home Bay | Kericho | Kasama | Mporokoso | Siraha | Sakteng | Udzorong |
| Geographic features | Rocky as well as collapsible soils; High water table<br>Suba South: Near lake | Rocky and collapsible soil; Accessible roads | Clay and sandy soil; Frequent rain; Limited space (clustered houses) | Heavy clay soil; Fertile; Heavy rains | Fertile; Moderate weather; Occasional flooding | High altitude; Clustered houses; Cold climate | Steep hills; Moderate weather |
| Population density (remote, rural, peri-urban) | Rural or peri-urban (depending on the village) | Rural or peri-urban (depending on the village); Small hills | Peri-urban; Near a large town | Lunte: Remote, very rural with poor road network | Peri-urban; Rural | Rural; Accessible roads to town centers | Rural; Villages connected through farm roads |
| Livelihood | Farming; cattle; Fishing | Coffee; Tea; Farming on plantations | Small businesses; Selling in markets | Chishamwamba: Farming compounds | Farming; Animal husbandry | Animal husbandry; Migratory populations | Subsistence farming; Cattle |

### 2.2. Data collection

Qualitative data, including KIIs and FGDs, was collected from November 2019 to March 2020 in Kenya, Zambia, and Nepal at the national, sub-national, and community levels. Data was collected in Bhutan in December 2020, due to delays from COVID-19; however, we did not observe any impact on sustainability of sanitation coverage in Bhutan as a result of the COVID-19 pandemic. Data collection activities are summarized in Table 2.

**Table 2.** Summary of qualitative data collection activities and participant breakdown.

|  | **Kenya** | **Zambia** | **Nepal** | **Bhutan** | **Total** |
| --- | --- | --- | --- | --- | --- |
| **Interviews**<br>*Key: N of KIIs (Participants)* | 18 (22) | 18 (22) | 23 (23) | 13 (13) | 61 (80) |
| SNV Staff | 4 | 2 | 2 | 2 | 10 |
| National Govt | 2 | 1 | 2 | 2 | 7 |
| District Gov't | 3 | 5 | 3 | - | 11 |
| Sub-district Gov't | 9 | 3 | 4 | 4 | 20 |
| Village | 1 | 6 | 7 | 2 | 16 |
| Local NGO | - | - | 4 | - | 4 |
| Private Sector | 3 | 5 | 2 | 3 | 13 |
| **Focus groups**<br>*Key: N of FGDs* | 12 | 10 | 10 | 8 | 40 |
| FGDs with women | 6 | 5 | 5 | 4 | 20 |
| FGDs with men | 6 | 5 | 5 | 4 | 20 |
| **FGD participant breakdown**<br>*Key: N of Participants* | 89 | 98 | 82 | 79 | 348 |
| Elderly (55+ years old) | 25 | 29 | 15 | 19 | 88 |
| Persons with disability | 9 | 6 | 1 | 2 | 18 |

Interview guides were adjusted to local context with input from field staff.^24^ Guides were translated into local languages when necessary. When appropriate, interviews were conducted by the lead researcher in English. All community FGDs were held in local languages identified by field staff. National-level interviews were conducted with government officials and representatives from international organizations and development agencies. Sub-national-level interviews were conducted with public health officers, community health workers, masons, and community leaders. Interviews included one or two key informants and lasted 45-90 minutes. FGDs were conducted with community members at the village level. Each FGD included six to twelve participants and lasted about two hours.

With permission from participants, we audio-recorded KIIs and FGDs, except for some national-level interviews where we relied on notes for analysis. Recordings were transcribed and translated into English. All recordings and transcriptions were password-protected and uploaded to a secure folder. Debriefs with the lead researcher and all research assistants were completed daily to discuss emerging themes, follow-up questions, and potential iterations to the topic guides.

### 2.3. Tool development

Data collection tools, including topic guides and demographic surveys, were developed primarily by our research team. We collaborated with SNV in-country staff to ensure that topic guides sufficiently probed for previously identified barriers and enablers and to confirm that all data collection activities were culturally appropriate and acceptable.

#### Desk Review

To inform tool development and conceptual framing, we conducted a thorough desk review of SNV program documents to further understand the SSH4A approach and completed a comprehensive literature review of rural sanitation methodologies. We also evaluated key findings from SNV’s performance monitoring data which measured improvements in sustainable programming.^21, 24^ Table 3 outlines the four components of the SSH4A approach, expected outcomes, and indicators to measure success and sustainability throughout implementation.

**Table 3.**
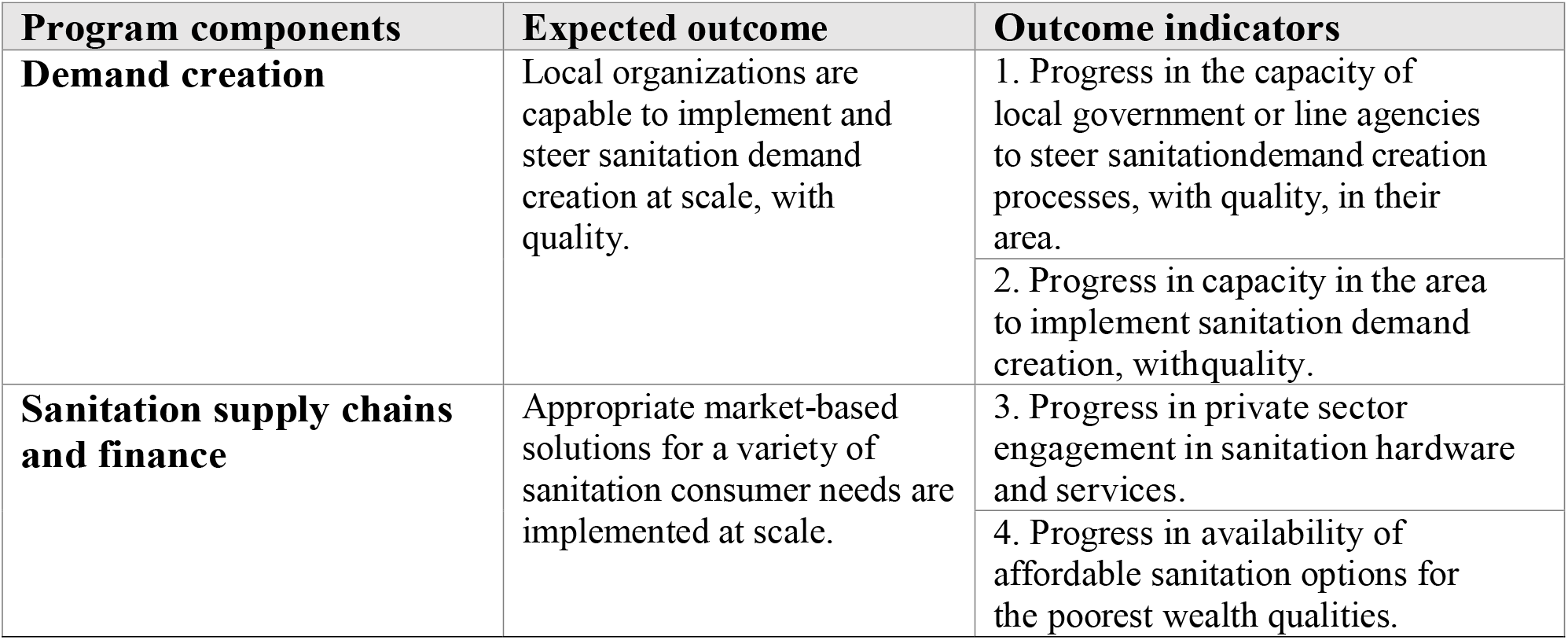
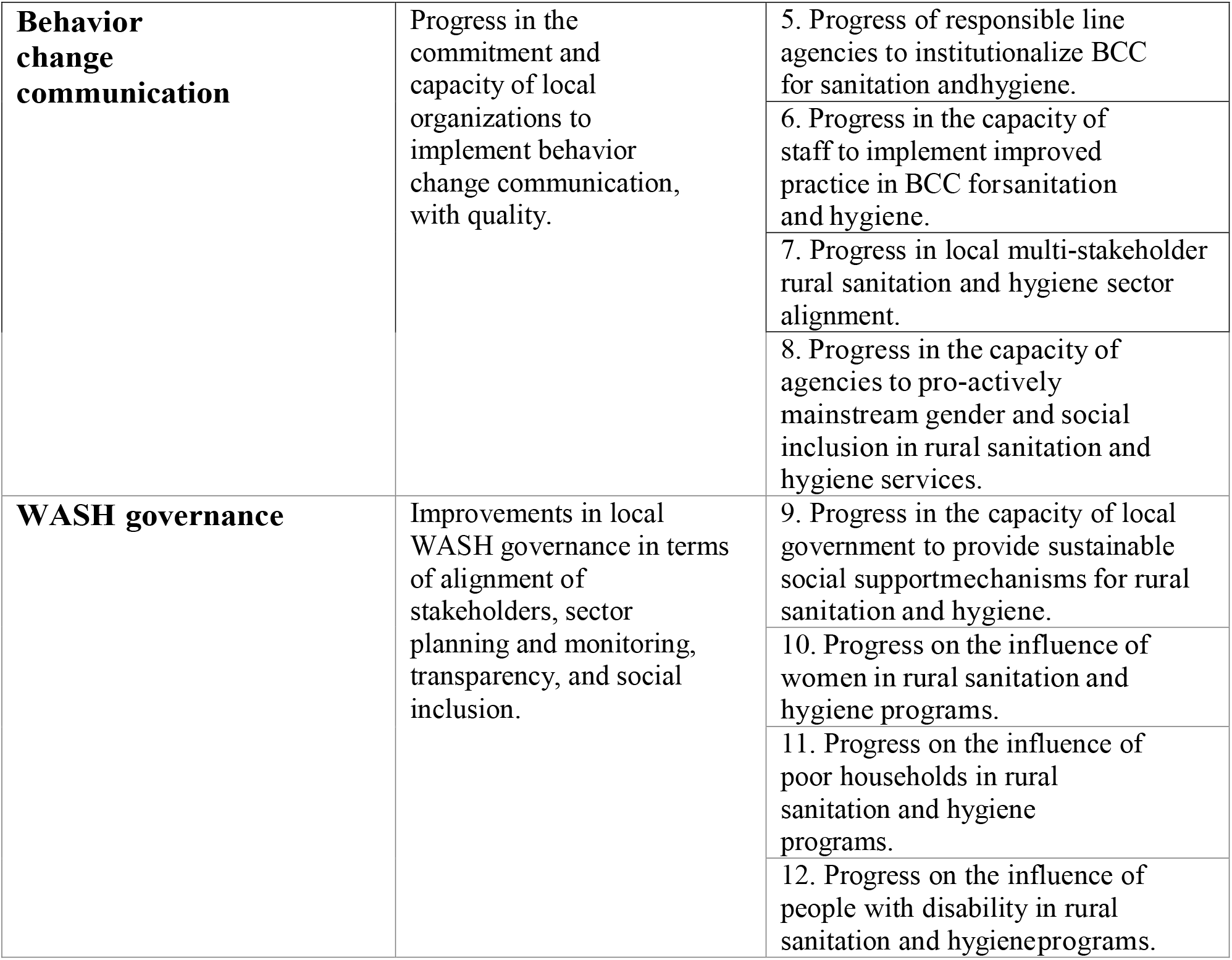
SSH4A list of outcome indicators.

| **Program components** | **Expected outcome** | **Outcome indicators** |
| --- | --- | --- |
| **Demand creation** | Local organizations are capable to implement and steer sanitation demand creation at scale, with quality. | 1. Progress in the capacity of local government or line agencies to steer sanitation demand creation processes, with quality, in their area. |
|  |  | 2. Progress in capacity in the area to implement sanitation demand creation, with quality. |
| **Sanitation supply chains and finance** | Appropriate market-based solutions for a variety of sanitation consumer needs are implemented at scale. | 3. Progress in private sector engagement in sanitation hardware and services. |
|  |  | 4. Progress in availability of affordable sanitation options for the poorest wealth qualities. |
| **Behavior change communication** | Progress in the commitment and capacity of local organizations to implement behavior change communication, with quality. | 5. Progress of responsible line agencies to institutionalize BCC for sanitation and hygiene. |
|  |  | 6. Progress in the capacity of staff to implement improved practice in BCC for sanitation and hygiene. |
|  |  | 7. Progress in local multi-stakeholder rural sanitation and hygiene sector alignment. |
|  |  | 8. Progress in the capacity of agencies to pro-actively mainstream gender and social inclusion in rural sanitation and hygiene services. |
| **WASH governance** | Improvements in local WASH governance in terms of alignment of stakeholders, sector planning and monitoring, transparency, and social inclusion. | 9. Progress in the capacity of local government to provide sustainable social support mechanisms for rural sanitation and hygiene. |
|  |  | 10. Progress on the influence of women in rural sanitation and hygiene programs. |
|  |  | 11. Progress on the influence of poor households in rural sanitation and hygiene programs. |
|  |  | 12. Progress on the influence of people with disability in rural sanitation and hygiene programs. |

#### Multi-country workshop

Prior to formal data collection, we held a workshop with program managers, implementors, and stakeholders at the SNV learning event in Accra, Ghana in August 2019. Program staff from 11 countries participated in this activity (countries included: Ghana, Burkina Faso, Ethiopia, Kenya, Uganda, Tanzania, Rwanda, Zambia, Nepal, Bhutan, and Laos). Together, we reconstructed the SSH4A implementation process to highlight program components, outputs, and outcomes across a variety of contexts. The purpose of this activity was to understand the SSH4A approach from the implementers’ perspectives. As KIIs and FGDs were held 1-2 years post-intervention, it was important to collaborate with SNV staff in order to understand what activities were conducted through the external intervention. The implementation process flow chart, illustrated in Figure 1, provides further details about the activities conducted or supported by SNV. Data collection and analysis tools, including topic guides and codebooks, were informed by the information gained from this workshop as we probed participants to discuss how capacity building activities implemented by SNV strengthened service delivery, and to identify gaps post-intervention. Key themes, activities, and potential sustainability factors that emerged from this workshop were introduced through probes in topic guides and as codes in analysis.

**Figure 1.**
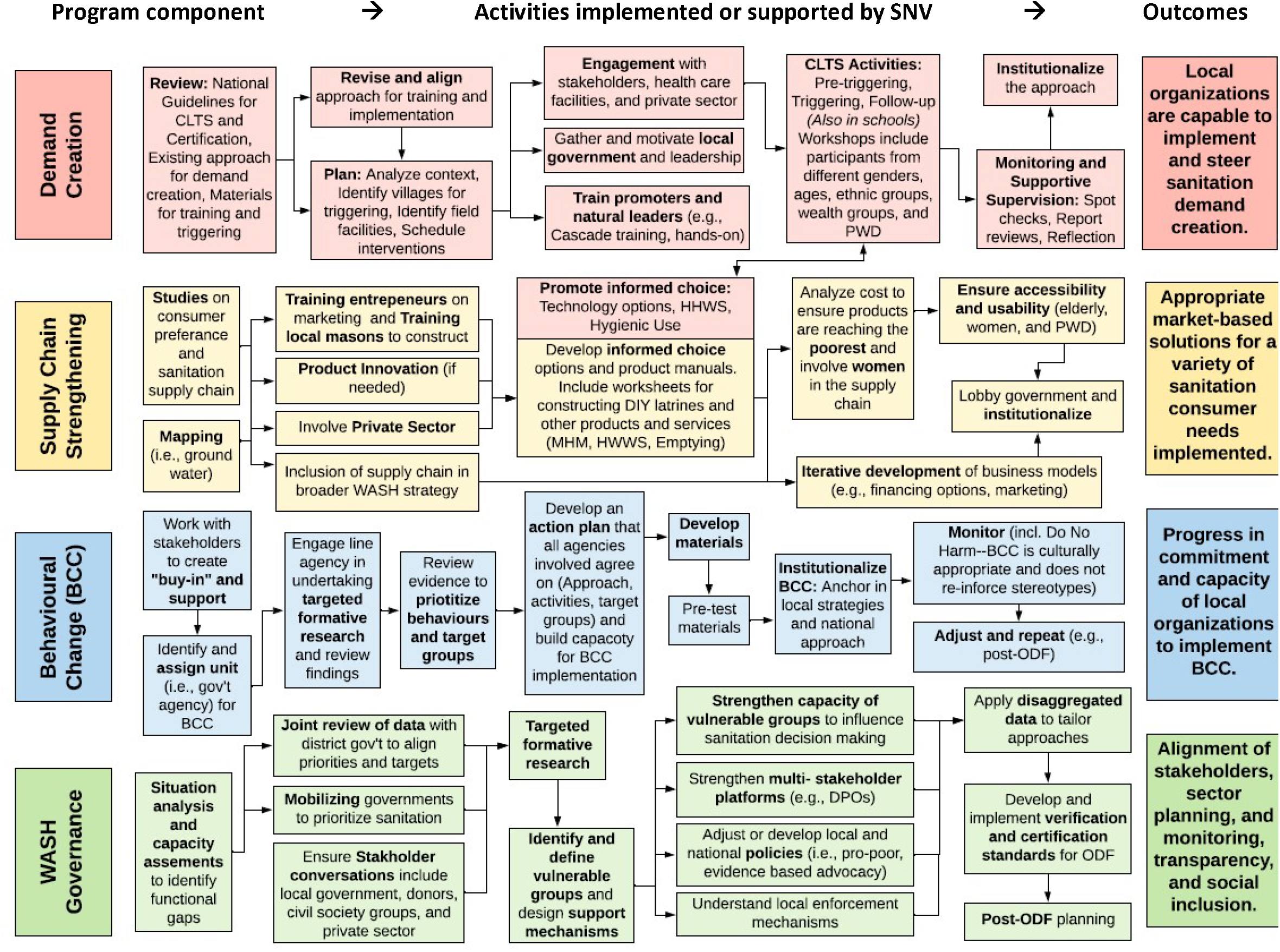
SSH4A programmatic implementation process flowchart reconstructed by SNV program staff from 11 countries in Africa and Asia.

### 2.4. Qualitative data analysis

We conducted inductive and deductive thematic analysis to identify factors related to the sustainability of sanitation improvements. The qualitative codebook was developed using a deductive approach, applying ideas from existing rural sanitation frameworks, which focused on: institutionalization of services, policies, governance, capacity building, financing, infrastructure, and monitoring. The Ottawa Charter for Sanitation Stages (OCSS) framework combines service delivery factors (e.g., affordability of latrines, acceptability of messaging) with behavioral, environmental, cultural, and structural factors related to sustainable sanitation.^25^ We primarily applied factors from the OCSS framework, with additions from other frameworks as needed to appropriately categorize the data. Inductive codes were added to account for emerging themes. All analyses were conducted using MAXQDA software. The data analysis process and codebook are outlined in Figure 2 and Table 4 (the fifth column in Figure 2, titled “coding”, corresponds with codebook described in Table 4). The comprehensive codebook and topic guides for KIIs and FGDs are available on our OSF page.^24^

**Figure 2.**
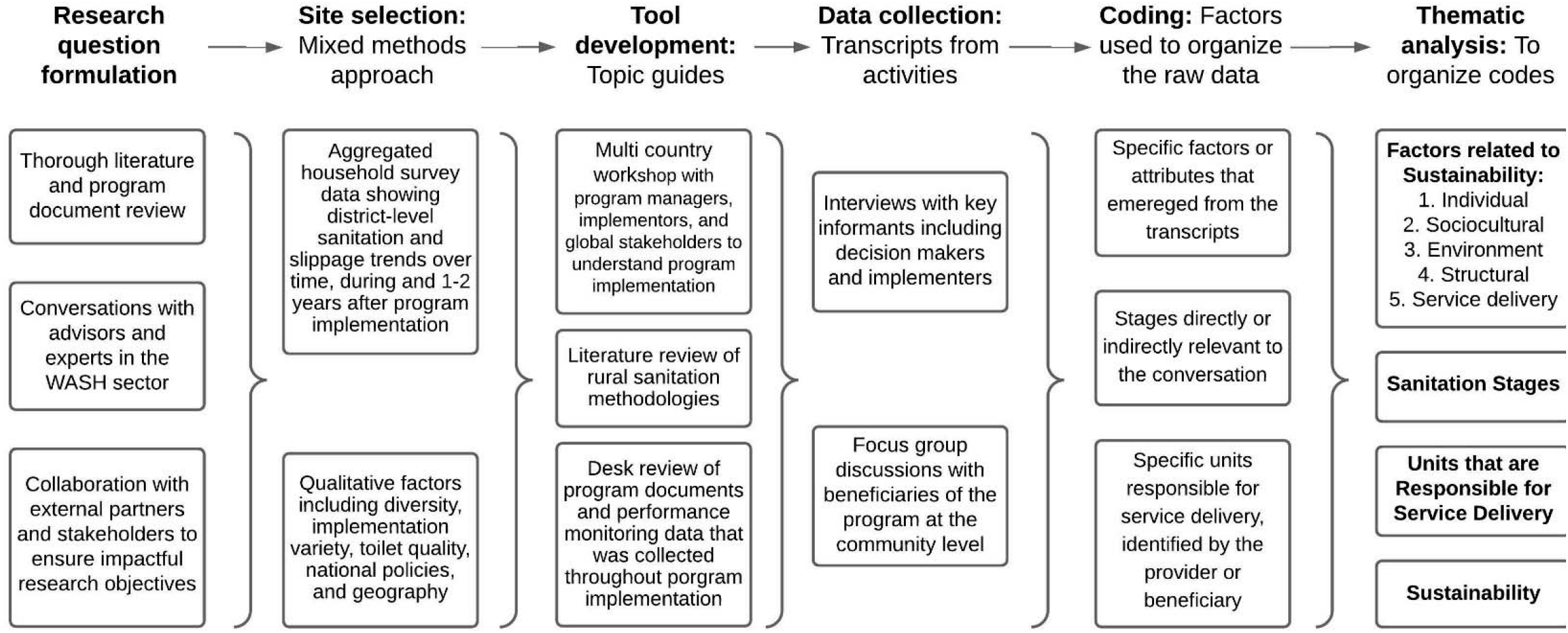
Data analysis process and definitions of key terms for qualitative research.

**Table 4.**
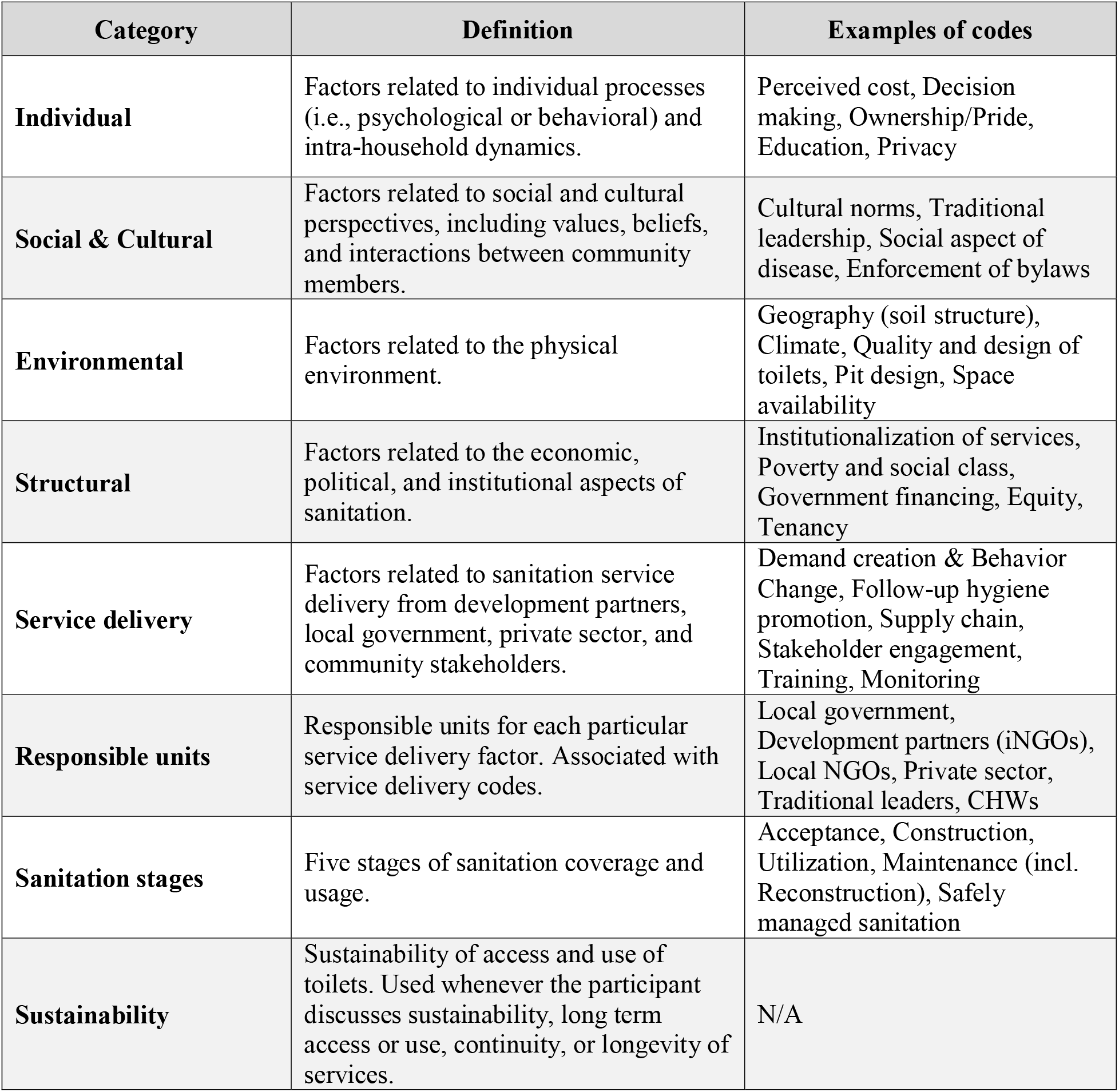
Summary of the codebook used for analysis, adapted from OSCC framework ^25^.

## 3. Results

### Summary of sanitation coverage and sustainability across four case studies

Rural sanitation improvements were generally sustained in Kenya, Nepal, and Bhutan, while there were higher levels of slippage in Zambia.^23^ Table 5 illustrates historical and current rural sanitation coverage and slippage levels in the selected case studies for this study. In Zambia, because of the large population size and significant differences between sub-district areas (e.g., wards), we conducted an additional ward-level analysis to ensure qualitative data was collected in diverse locations (not shown in table). We found that there was no recorded slippage in Chishamwamba (+4%) and elevated slippage in Lunte (−17%), two wards in Mporokoso; and high levels of slippage in Chiba (−61%) and Lua-Luo (−52%), two wards in Kasama. These localities are referred to in the qualitative results.

**Table 5.**
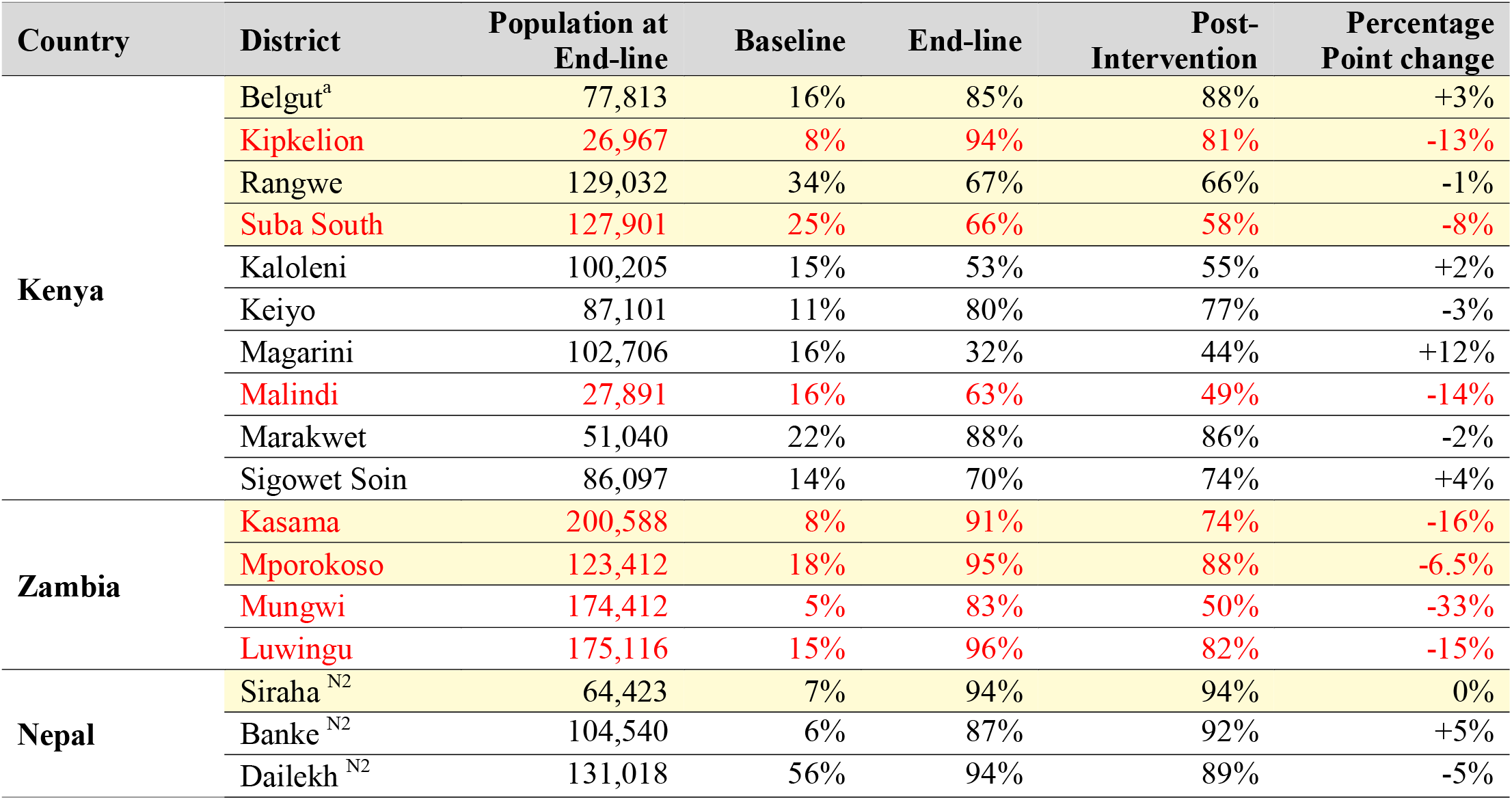
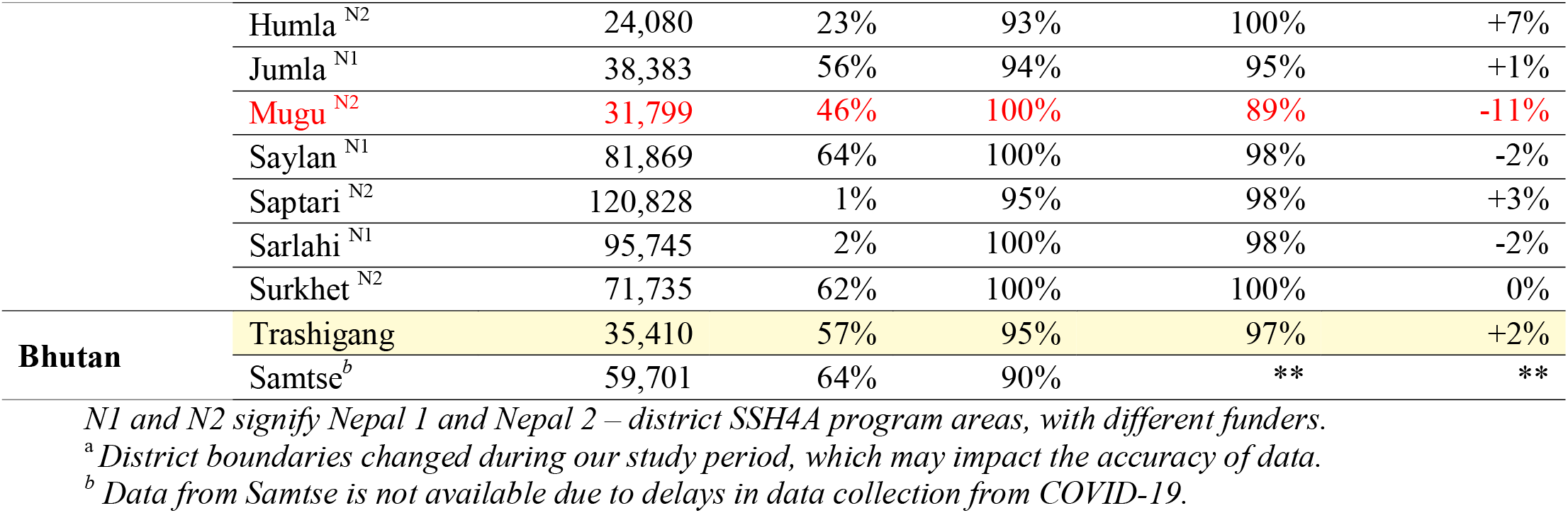
Sanitation coverage and slippage rates for SSH4A program areas. Highlighted rows were selected for qualitative research. Districts with elevated slippage are in red. ^23^.

### Sustainability factors

The findings presented in this section were derived from analysis of data from KIIs and FGDs, which were conducted 1-2 years post-intervention. The heat map below, illustrated in Table 6, compares sustainability factors across countries assessed in this study. The sustainability factors are separated into categories: 1) Service delivery, which refers to components of the local service delivery system; 2) Contextual, which refers to environmental and social factors; and 3) Behavioral, which refers to individual factors related to decision-making. Sustainability factors can be described as barriers (hindering sustainability) or enablers (driving sustainability). For example, in Kenya and Zambia, lack of *access to materials* was typically discussed as a barrier to sustainability, while in Nepal and Bhutan easy *access to materials* was discussed as an enabler. In all four countries, *geographical factors* were discussed as either barriers or enablers, depending on the location within the country – areas with collapsible soil or frequent flooding discussed barriers, while areas with flat land and clay-like soil for brick construction discussed enablers.

**Table 6.**
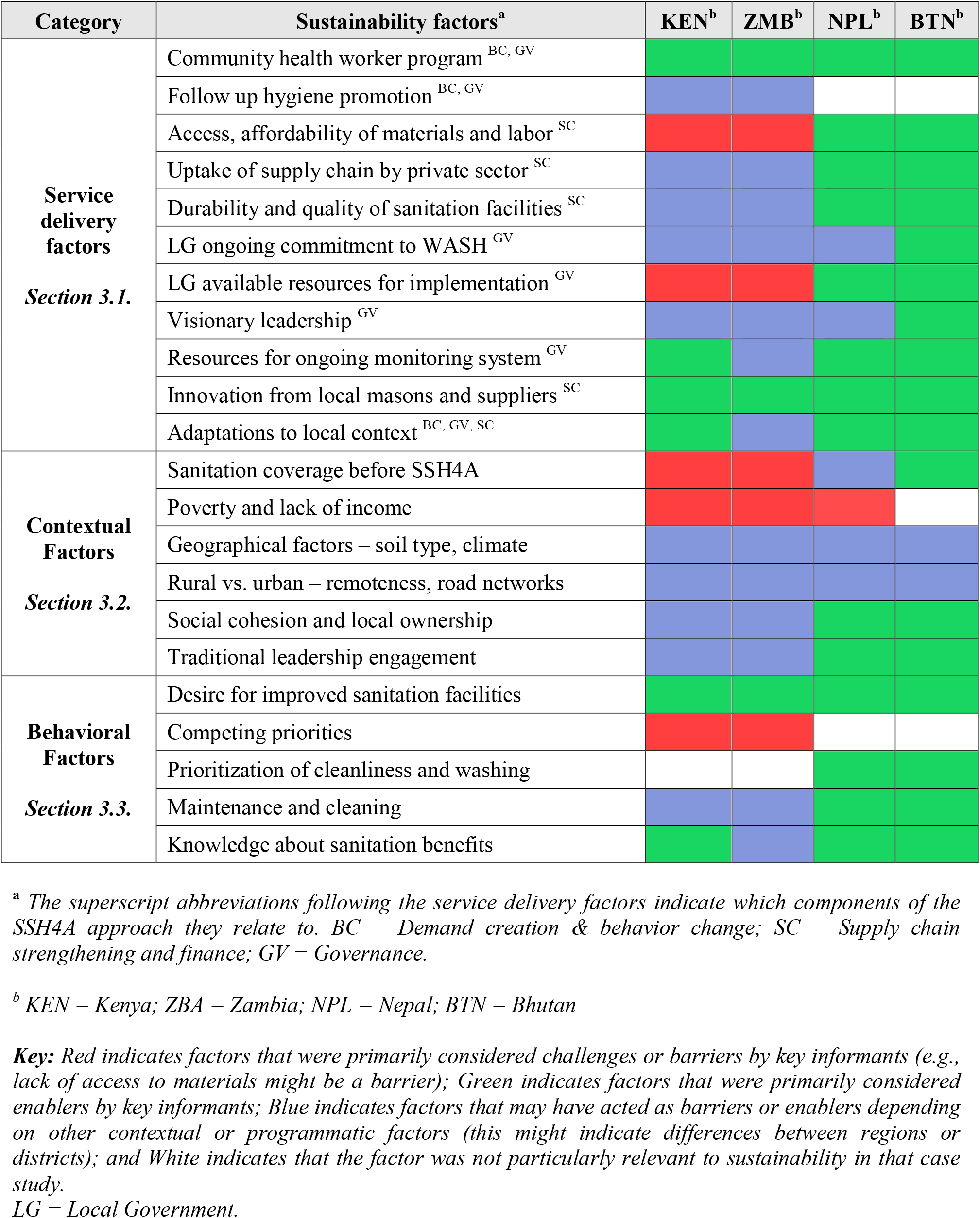
Heat map illustrating sustainability factors for rural sanitation coverage in Kenya, Zambia, Nepal, and Bhutan.

### 3.1. Service delivery sustainability factors

Service delivery sustainability factors represent characteristics or components of the local service delivery system 1-2 years post-intervention, as discussed by key informants interviewed in this study. Factors, illustrated in Figure 3, are separated into categories that emerged from the data: 1) demand creation and behavior change; 2) supply chain and finance; 3) governance; 4) capacity building; 5) monitoring and evaluation; 6) adaptive programming.

**Figure 3.**
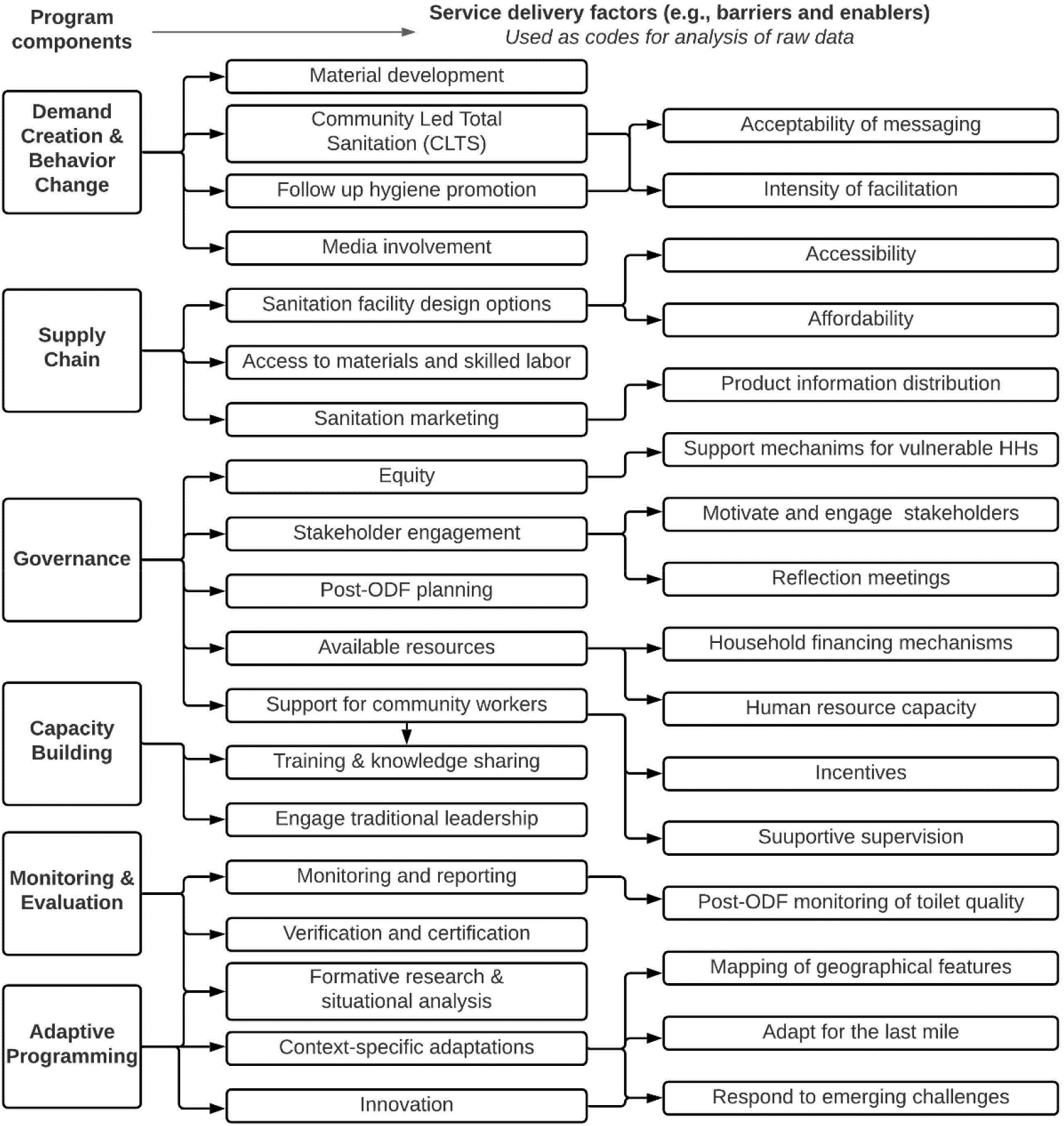
Service delivery factors: Coding tree from analysis of raw data.

#### Demand creation and behavior change

Community health worker programs were primarily described as enablers for sustainability as they represent local capacity for implementing demand generation and behavior change activities. The ability of community workers to adapt to social and cultural norms relevant in the areas where they work and live may also support sustainable programming. SNV, with support from local governments, typically led trainings for community workers, volunteers, and traditional leaders.

> *“The most important thing is door-to-door outreach, where you talk to people at individual level to understand why they don’t have a latrine, or why they have a latrine but they are not using them.” (SNV staff, Kenya)*

Continuous follow-up for health promotion was also discussed as an essential aspect of sustainable service delivery. Consistently reminding community members about the importance of latrine construction, use, and maintenance may be required months, even years, after a community is open defecation free. Additionally, tailored messaging for households who need to reconstruct latrines after they have collapsed likely helped to maintain coverage in areas where lower latrine quality was more common.

> *“People want to see us many times. That is when they realize it is serious. If you sensitize them and then you forget about them, they also forget about the whole thing. When they see us often, it sticks more.” (CLTS coach, Zambia)*

#### Supply chain and finance

Access to materials (e.g., plastic, cement, culverts, bricks) and skilled labor varied between and within countries, and was identified as a sustainability factor by all key informants (n=80). In Kenya and Zambia, lack of access to materials was commonly mentioned as a challenge for constructing durable latrines. This lack of access may be due to affordability of materials, poverty and lack of income, or poor road networks to central urban locations.

> *“You will find that lack of finances to buy [cement] leads people to build the other pit latrines that we have been talking about that collapse in [less than] two years. We don’t have enough resources to build permanent latrines that won’t collapse.” (Woman, Lua-Luo, Kasama)*

However, most FGD participants also discussed the use of locally available materials, including logs and bricks made from clay-like soil, which were more accessible.

> *“Most of the toilets here, when we build them they last many years and the reason why they last many years is because the soil here is like clay. Therefore, the toilets do not collapse inside so the latrines last very long because the soil is strong.” (Community leader, Chishamwamba, Zambia)*

In Nepal and Bhutan, these issues were not as prevalent, and access to affordable materials for durable latrine construction was considered an enabler in some areas. For example, in Bhutan, the Forest Park Office (a government entity) provided free timber for sanitation facility construction.

> *“When people raised the concern of not having access to timber to build toilets, we also had a strategy for close collaboration with the Forest Park Office to allocate free timber to every household, which facilitated the construction of toilets. I think the results we achieved have been mainly due to close collaboration and teamwork between [local] leaders, the Ministry of Health, and the Forest Park Office.” (Community leader, Bhutan)*

Key informants, especially district level government staff and active masons, also discussed uptake of supply chain improvements by the private sector. In all program areas, key informants reported that SNV trained masons to construct toilets, adapt construction and design to account for geographical factors and individual preferences, and market their products to consumers.

> *“SNV once brought a project to build toilets with masons, so they discussed with us how these toilets will be built and we had different prices for these toilets based on how much one could manage to pay… I personally joined this group.” (Woman, Lua-Luo, Zambia)*

During training, SNV also prioritized accessibility of toilet design for the elderly and persons with disabilities. This included hand rails and raised toilet seats.

> *“SNV has also made a manual about how to make disabled friendly toilets with options. We talked about it during mason training” (Local NGO staff, Nepal)*

In Zambia and Kenya, 1-2 years after SSH4A implementation was complete, masons reported that they were working at the district level (often headquartered in the main town) and traveled to many communities for work, making very remote areas unappealing for business. One mason from a remote community, who was trained by SNV, claimed he was unable to work as a mason because he could not access the required materials. The village he lived in was on a dirt road that often flooded and was over three hours away, driving, from the closest mid-sized town. In Bhutan, where collaboration between SNV and the national government was particularly seamless, these trainings were led by the local health sector with support from SNV staff. Although the success of implementation varied between case studies, all of the masons that were interviewed (n=13), in all four countries, discussed the knowledge and skills gained from training led by SNV.

> *“The government health sector invited me to a meeting and that gave me the opportunity to be involved in sanitation business. In town, there was already an entrepreneur who was doing sanitation business. And there was no one running a sanitation business in my [rural] area, despite having a bigger population. The officials asked me if I could be a supplier… I agreed to supply all kinds of materials like toilet pots, pans, pipes, tin sheets, and even the plumbing equipment. This initiative from the government helped me a lot in my business and even benefitted the households as people do not have to travel to far places to buy sanitation materials.” (Small-Medium Enterprise, Bhutan)*

Additional sanitation marketing activities, including distribution of product manuals to promote informed choice of facilities, advertisement of improved toilets in schools and public spaces, and consumer research to identify preferences, were also mentioned by key informants. However, sanitation marketing initiatives diminished in some areas, according to participants, without continuous support from SNV.

> *“If somebody wants to have their latrine improved, [SNV] had formed a [sanitation marketing] committee [which would assist with in-kind payment options]. But when they went to the local authority to get the [materials] they use because of the transfers, it couldn’t work out because today they talk to one person, and then they were transferred, and so it became so difficult.” (Woman, Kasama, Zambia)*

Strength of the existing supply chain, uptake of supply chain improvements by the private sector, affordability of materials and labor, and socioeconomic factors all had an impact on the durability and quality of sanitation facilities. In Kenya and Zambia, some areas had higher quality latrines (Belgut, Rangwe, Chishamwamba), while others had very poor-quality latrines – commonly referred to as “temporary” latrines (Suba South, Kipkelion, Kasama). Participants from communities with lower-quality latrines discussed the difficulties associated with frequent reconstruction – including finances, time, and lack of motivation to reconstruct.

> *“If you visit a household and they don’t have a latrine, you will ask them to build it using locally available materials, but as time goes maybe the locally available material actually fades, and they have not changed them into a better latrine, so they will stop using it especially the children because they will fear they could fall inside” (Local government staff, Belgut, Kericho)*

In Nepal and Bhutan, most latrines were durable, regardless of climate, as they were made from strong materials. In Nepal, participants in all FGDs (n=10) reported that no one in their community needed to reconstruct, and described that all toilets in their area were built with either cement rings (e.g., culverts) or septic tanks.

> *“There are four or five types of toilets. Pit latrines are one option, but they are not safe. Mainly, we can build two kinds of toilets: toilets with septic tanks and toilets with cement rings are found in our community.” (Woman, Mahadeva, Nepal)*

#### Governance, capacity building, and monitoring and evaluation

According to key informants at district, regional, and national-levels in all four countries, the SSH4A approach invested considerable time and resources into strengthening sustainable governance structures through capacity building and technical support. However, the extent to which local governments maintained sufficient financial and human resources for sanitation activities varied between districts and countries. Ongoing commitment from local governments after SSH4A implementation was commonly cited as an important sustainability factor, and areas that lacked local government commitment and resources might have experienced more slippage. However, there was a clear distinction between commitment from the implementation level (e.g., district, ward, or municipality offices) and commitment from the national-level. Especially in Kenya and Zambia, there were health officers at the district or ward level that were highly committed to improving sanitation in their area, but the lack of national-level government commitment resulted in a lack of financial and human resource allocation to the district officers for sanitation activities.

> *“I think there was a bit of relaxation during SNV’s handover, such that things couldn’t continue like the way they were done previously, at the pace it started with. What is sad is that, earlier on, even up till now, when the program was being conducted, not all clinic facilities had environmental health technologists. The Environmental Health Technicians [EHTs] are responsible for the continuation of sanitation activities in catchment areas…For it to continue, there is a need for capacity building of the EHT’s who are in those health facilities so that they continue with the behavior change activities.” (Environmental Health Officer, Zambia)*

Local governance systems were also augmented by post-ODF planning (e.g., sustainable behavior change, maintenance, and fecal sludge management), continuous training and engagement with community workers, and attention to equity. Data also suggested that community members who were aware of their local government’s commitment to sanitation were more inclined to prioritize sanitation themselves. Related is the local governments’ ability to support continuous training and engagement for community workers, traditional leaders, and new staff members, and to support monitoring and evaluation activities once external funding and support concludes.

> *“There was a royal visit to Sakteng, and His Majesty the King said that he will not except anything but good sanitation, hygiene, and health from all of us, the people of Sakteng… this has inspired and motivated us to improve our sanitation and hygiene. Since then, every month, we have met and started improving. And the local health assistants, park officials, and even police officers continue to follow-up and support identifying locations for toilets and provide other technical advice.” (Man, Sakteng, Bhutan)*

#### Adaptive programming

The capacity of local governments, private sectors, and local non-profit organizations to adapt or alter programmatic activities and sanitation service delivery after SSH4A implementation was completed varied between program areas. Adaptive capacity may be affected by training facilitated by the development partner, resources allocated for reflection and evaluation, the presence or availability of people who have the skills needed to innovate and respond to challenges, and employee turnover in local health offices. Two components of adaptability that were most frequently mentioned by key informants were the ability for local masons to innovate on toilet designs, and the ability for local decision-makers to adapt to contextual changes or specific needs. For example, in areas with masons who were innovative and could adapt toilet design to withstand geographical challenges, sustainability of individual latrines was more common.

### 3.2. Contextual sustainability factors

Contextual sustainability factors are not direct objectives of sanitation interventions or activities, and may not be able to be altered or improved through WASH interventions, but are still identified by key informants as being related to the sustainability of sanitation improvements. These factors are usually considered when designing sanitation programs because adaptations or innovations may be required in order to address contextual barriers or enablers. For example, in areas with thick, clay-like soil, community health workers may teach individuals how to build their own superstructures out of locally made bricks. However, in areas with collapsible soil, supply chain strengthening to support the development of sanitation technologies that are both durable and affordable may be more appropriate.

Table 7 summarizes the influence of contextual factors on sanitation service delivery – the first column lists contextual sustainability factors, and the second column describes how these contextual factors may have a positive, negative, or neutral impact on service delivery factors identified by key informants. We discuss each of these contextual factors individually in the sections below.

**Table 7.** Impact of contextual factors on sanitation service delivery.

| Contextual factor | Impact on service delivery factors |
| --- | --- |
| Poverty, income | Latrine options, household financing, subsidies |
| Gender roles | Maintenance responsibility, target for demand creation |
| Marginalized groups, and defiant groups | Tailored messaging, enforcement from traditional leaders or local government |
| Soil type, climate | Supply chain, toilet design, sanitation marketing, messaging for reconstruction, monitoring consistency |
| Rural vs. urban, remoteness | Tailored messaging to adapt to cultural differences, accessibility of materials, toilet design, tenancy considerations, how fecal sludge is safely managed |
| Space availability | Toilet location in relation to the house, quality and durability, how fecal sludge is safely managed |
| Social cohesion and values | Demand creation activities, tailored messaging |

#### Poverty and lack of income

Poverty was identified as one of the most pressing concerns in regards to the sustainability of sanitation coverage from the perspectives of our key informants, especially in Kenya and Zambia. Lack of income (in some areas, lack of any available cash) prevented community members from investing in durable latrine options because they could not afford high-quality materials (e.g., cement, culverts, bricks) or the labor required to dig strong, deep pits that would not collapse or fill up quickly.

> *“People love durable toilets, but fail to raise the money to pay [for them]. People appreciate the improved kinds of toilets that we build, it’s just that they don’t have money to afford.” (Mason, Zambia)*

Consequently, lack of affordable high-quality materials may also be due to underdeveloped markets, especially in African countries, which further exasperates this inequity. The cost of cement, slabs, and culverts in Zambia and Kenya was considered expensive by most participants.

> *“Financial problems can be a challenge, I want to build [my latrine] with cement, I want to buy a slab for it, but these other temporary latrines, we just build them because we don’t have income [to pay for durable materials] … if income was here, we could construct the long-lasting ones, me that’s my opinion.” (Woman, Rangwe, Homa Bay)*

In most areas, formal household financing mechanisms were scarce or unavailable. However, community savings groups and in-kind payments to masons offered an opportunity for households in the lowest wealth quintiles to invest in sanitation facilities.

> *“You can get some money from a savings group, then it becomes a loan because you have to pay back with interest. So as a community, we do not get loans from the bank, but from savings groups.” (Man, Chishamwamba, Zambia)*

In Nepal, communities who self-identified as poor still claimed they would construct “good toilets” through loans or support from the community, and that they would construct high-quality toilets even though they were poor. Overall, community members in Nepal reported that open defecation was “not an option” regardless of socioeconomic challenges.

> *“Poor people who have no money and who have no one in their household to make money, they have taken loans out to build toilets, because there is no option of building a toilet. One must make toilet in our community.” (Man, Nepal)*

#### Geographical and environmental challenges

Geographical barriers, including sandy or rocky soil, high water table, heavy rain, and flooding were mentioned in all counties, but not in all program areas. Typically, these barriers seemed more intrusive to participants who used lower quality latrines rather than durable toilets.

> *“Slippage happens during rainy season when the floods happen, most of the toilets collapse and coverage goes down and people need to start [building a latrine] again.” (County government staff, Homa Bay, Kenya)*

Innovations were applied to adjust to these geographical barriers, including raised toilet platforms to avoid flooding pits and building toilets inside to prevent destruction from winds. However, innovations were more common in areas with durable latrines, because use of locally available materials does not provide as much flexibility in terms of facility design.

> *“There are toilets which are built outside of houses. Weather could affect and become a problem to these households. There is risk of the roofs being blown off by the wind storm, collapsing of walls and breakage of toilet pots from the falling stones from the rooftops.” (Mason, Bhutan)*

Sometimes preventing damage to toilets was not possible, and households would need to reconstruct. Barriers for reconstruction included available finances, time, and motivation. Soil type was particularly important for communities that relied on locally available materials for household constructed pit latrines.

> *“Because of the soil we have, latrines collapse, these trenches that we dig, when water goes in there, it collapses. We will thus start looking for another place to build another toilet and so we are running out of land”. (Woman, Lua-Luo, Kasama, Zambia)*

#### Community ownership and social cohesion

Key informants also described the importance of social cohesion and knowledge of the social nature of disease transmission (i.e., if one household does not have a latrine, it impacts the whole community) in relation to demand generation activities targeted to rural households.

> *“OD [open defecation] practices indirectly affect those with latrines because flies will feed on the feces and land on the meal of those who own latrines. So, those who have latrines must continue to motivate people practicing OD to build a latrine, because sometimes, children drink water directly from the dam which might be contaminated with feces, and this causes waterborne diseases in the community. When people drink that water, they can get cholera or diarrhea, and you know that when cholera infects 10 people in a family, it’s very difficult for them to survive and therefore that’s why we must strive to motivate and educate people on the importance of latrine usage.” (Man, Homa Bay, Kenya)*

Education for community members on disease transmission was supported by the palpable decrease in disease prevalence overtime as community members witnessed improvements in sanitation coverage – allowing for a tangible understanding of the importance of constructing and using latrines. Additionally, collective responsibility, social cohesion, and altruism within communities and between neighbors supported ownership of sanitation coverage at the village level and promoted support for vulnerable individuals in some program areas.

> *“The most important thing is for them to understand that they are responsible for their health. To maintain sustainability, we try to be there to ensure that whatever we did in the initial phase is maintained but I can’t say that it’s not challenging, it’s challenging.” (Local government staff, Kericho, Kenya)*

There were definitive differences between contextual factors, including social cohesion, even within countries. For example, social cohesion is generally a very effective facilitator for sanitation programming in African countries, including Kenya and Zambia. Most key informants in both countries reported that community members often help their vulnerable neighbors construct latrines. However, select areas visited for this study, including several wards in Kasama district in Zambia, lacked social cohesion and effective traditional leadership due to their peri-urban nature and rapid population growth as emerging town centers.

> *“For the rural communities, if we talk about issues of money - some are not able to access money easily. So, the only way they can do that is through my earlier point when I said that the communities themselves should mobilize, once they mobilize themselves, they set laws – the headman should say ‘We all need to have [high-quality] toilets in my community!’ Then, they sit down and they can help each other.” (Local government staff, Kasama, Zambia)*

### 3.3. Behavioral sustainability factors around sanitation uptake

Behavioral sustainability factors were identified by key informants, especially beneficiaries at the community level, as barriers or enablers for individual or household level decision-making related to the construction or use of sanitation facilities. These considerations likely impacted the uptake of sanitation service delivery and were often tied to social or cultural values and norms. Some behavioral sustainability factors included a desire for improved latrines, competing priorities, cultural values of cleanliness, gender roles related to maintenance and cleaning, and knowledge and education of sanitation benefits. Key informants noted that these behavioral factors play an important role in the sustainability of sanitation improvements, and should be considered when designing follow-up messaging and sustainable programming.

## 4. Discussion

Sustaining sanitation coverage, defined as access and use of household sanitation facilities, remains a critical challenge in the WASH sector.^6, 26^ This study focused on the identification of factors that likely contributed to the sustainability of sanitation improvements in four countries, and described how these factors may relate to varying levels of slippage 1-2 years post-intervention.^23^ Overall, the sustainability factors identified through this research aligned with findings from previous studies.^16–18, 20, 26–29^ To build on existing literature, we also explored the extent to which these factors either facilitated or hindered sustainability within four case studies, and how service delivery systems either utilized or addressed the factors based on context. The presence or absence of sustainability factors may have implications on where certain programmatic approaches will work, and where adaptations may be required.

According to our data and previous studies, one of the main challenges to achieving sustained sanitation is household poverty – considering both lack of income and lack of affordable sanitation options – which prevents households from constructing high-quality and durable sanitation facilities due to the high cost of materials (e.g., bricks, culverts, cement) and skilled labor.^26, 27^ Additionally, heavy rains and flooding will frequently result in the collapse of low-quality toilets in areas where households do not have the financial ability to reconstruct, and individuals may lose motivation to prioritize sanitation as a result. However, these barriers may be overcome with application of interventions targeting social cohesion and sustained behavior change.^27–30^ We found that in communities with stronger social cohesion, households experiencing poverty reported that their neighbors, community savings groups, religious institutions, or family members would help them construct sanitation facilities. While WASH tend to focus on behavioral and individual factors relevant for household-level decision making, studies have shown that communities are often a more critical point of investigation.^31^ The impact of poverty on the sustainability of sanitation improvements was also identified in a concurrent analysis of household survey data which reported that households in the lowest two wealth quintiles had higher slippage rates.^23^ Lack of income was less problematic in select areas where community members had access to income through fishing, coffee and tea plantations, or farming compounds, some of which were also closer to urban town centers which had a more accessible supply of affordable materials.

In all program areas, a solid foundation for sanitation businesses was established; however, uptake of supply chain improvements by the private sector varied. Supply chain activities completed in all four case studies aligned with suggestions from existing literature, including mapping of supply chain actors, consumer profiling and willingness-to-pay studies, technical training for masons on improved latrine design, and provision of informed choice materials.^19, 32–34^ In Kenya, one product, called the SAFI latrine, was developed to address gaps in the supply chain after a consumer preference study revealed that appropriate latrine options were limited in the market.^35^ Sanitation businesses were slowly addressing this challenge, and improvements peaked in Homa Bay compared to other areas. In Zambia, sanitation marketing committees (referred to as SanMark) were established to support masons, suppliers, and consumers through training for construction, marketing, and self-financing as well as the development of activities and materials used to promote informed choice.^36^ However, in both Kenya and Zambia, where access to materials can be challenging, supply chain improvements seemed to diminish after SNV was no longer present to support capacity building. In Nepal and Bhutan, where access to materials was not mentioned as readily by participants, supply chain improvements were generally sustained by the private sector, and key informants could easily identify small business owners who supplied sanitation materials (e.g., plumbing, culverts, cement) in their communities.

Previous research suggests that geographical factors including climate (e.g., heavy rains), natural disasters, soil type, and terrain likely impact the sustainability of sanitation facilities.^17, 18, 27, 29^ However, we also found that the ability of the local service delivery system, including government officials, community volunteers, and masons, to adapt to geographical barriers also played a significant role. For example, in Nepal, participants mentioned the challenges of frequent flooding. To address this, masons constructed elevated toilets with doors and roofs that would not fill with water when it rained. Therefore, households were able to maintain their sanitation facilities regardless of flooding. In Suba South in Kenya, collapsible soil from being near Lake Victoria alongside frequent rains contributed to the frequent collapse of poor-quality pit latrines which likely hindered sustainability especially in low-income households, as uptake of the SAFI latrine had not yet peaked in this area.^35^

Population density, or the extent to which a village was considered remote, rural, or peri-urban, also impacted sustainability. In peri-urban villages in Kasama, Zambia, where slippage was highest, some of the approaches implemented through SSH4A (e.g., community-led) seemed no longer applicable as these areas became increasingly urban due to increases in tenancy, less space available for toilet construction, transient populations, and a general lack of community ownership.^27, 30, 37^ According to our data and supported by previous studies, social cohesion and effective local leadership typically support sustained open defecation free (ODF) status, especially through community-based behavior change interventions.^27, 30, 37^ We found that community-based demand creation activities were less effective in peri-urban areas where tenancy was common and households were close together but not necessarily reliant on each other, therefore lacking social cohesion and community ownership.

Lower density program areas – including Suba South, Lunte, and Trashigang – also encountered unique challenges that required adaptation, including lack of follow-up visits from health officers and difficulty reaching towns for materials.^15, 38^ Several solutions were identified, including strong traditional leadership in Lunte and improved farm road access in Trashigang. Migratory populations who work as cattle herders (Sakteng, Bhutan) or fishermen (Suba South, Kenya) also required innovative programming as they could not construct toilets while traveling for months at a time. In Suba South, health officers would contact traditional leaders in beach communities to ensure fishermen who were renting houses had access to latrines, and to ensure that their families had latrines in their home villages. Some leaders would allow workers to return home for one week to construct latrines for their wives and children.

According to analysis of household survey data, countries with higher levels of sanitation coverage and infrastructure at baseline were less likely to experience slippage of improvements.^23^ Similarly, program areas with large increases in sanitation coverage during the implementation period were more likely to experience slippage compare to those with marginal gains.^23^ According to key informants, the relatively mature sanitation service delivery systems in Asia were more likely to sustain improvements from SSH4A programming compared to the less mature systems in Africa.^25, 26^ For example, in Bhutan and Nepal (where there were higher levels of sanitation coverage at baseline, and accordingly very sustained coverage) respondents reported that they had been using sanitation facilities before the current project was implemented at scale. The investments in supply chain strengthening and promotion of durable toilets were more successful in Bhutan and Nepal where community members were already opposed to open defecation.^22^

Political leadership is considered more influential in driving decisions regarding sanitation service delivery compared to donors, development agencies, and international organizations.^15^ According to key informants in all four countries, the SSH4A approach invested considerable time and resources into strengthening sustainable governance structures through capacity building and technical support. However, the extent to which local governments maintained sufficient financial and human resources for sanitation activities varied between districts and countries. Local government commitment and resource allocation appeared to be more reliable in Nepal and Bhutan compared to Kenya and Zambia. In Nepal, we observed a social movement for sanitation, with all stakeholders prioritizing WASH improvements and motivated to reach ODF status nationwide. Additionally, declaring health as a human right (both in Nepal and Kenya) further strengthened the local government’s role as the sole duty-bearer for WASH programs. Collective responsibility among community members, and a unique partnership between the local government, local NGOs, and community volunteers led to steep and sustained improvements. In Bhutan, the relationship between the local government, especially at the national-level, and SNV was described as exceptionally strong, collaborative, and sustained. All key informants who worked with either the government or SNV mentioned the positive impact of this partnership on the sustainability of sanitation programming. The SSH4A approach is the official approach for sanitation policies within the government of Bhutan, and the national government works closely with SNV and other development partners, which has positive implications on policy development and funding.

We found that program areas with more sustainable sanitation coverage were described as having passionate and innovative local leadership, commitment from the local government, sufficient resources for continuous programming, and the ability to innovate and adapt to contextual challenges. Capacity building remained important for sanitation programming after the SSH4A program was complete, including training and engagement led by the local government. Our data suggests that adaptive capacity, which may be impacted by training led by development partner programming, resources allocated for reflection and evaluation, capacity to innovate and respond to challenges, and employee turnover in local health offices, was also related to the maturity of service delivery systems and consequently the sustainability of sanitation programming and coverage. Behavioral enablers, including a household member’s desire for improved sanitation facilities, understanding of the benefits of sanitation, and cultural norms supporting sanitation were found in most program areas, even those that experienced slippage. Even with sustained changes to key sanitation behaviors (which include sharing latrines, using poor quality latrines, and reconstructing after heavy rains), sanitation coverage after the end of project activities relied heavily on governance, supply chain activities, and contextual factors.

### Strengths and limitations

Strengths of our research approach included, 1) a wide-range of key informants from the community to national-level, which allowed for a comprehensive understanding of sustainability factors, and 2) continuous and extensive conversations and collaboration with SNV staff, which allowed for our analysis to reflect programmatic implications. An important limitation of this study was the timing of data collection. Interviews were conducted 1-2 years after implementation of the SSH4A approach was complete and there may be a lack of understanding or recall bias regarding what was actually implemented. Additionally, FGDs covered a small part of the sub-national areas within each case study, and components of the SSH4A approach peaked differently in different parts of the countries.

## 5. Programmatic Implications

Findings from our qualitative data, alongside extensive conversations with an advisory group and SNV global and in-country program staff, support several programmatic implications:

- The maturity of a country’s sanitation service delivery system will likely impact the timeline needed for programming from development partners. In some settings, longer timelines may be needed to strengthen less mature service delivery systems and to build the adaptive capacity in local governments and supply chains systems that is required to sustain sanitation service levels and maintain high coverage.
- The alignment and continuity of efforts to produce sustainable improvements in sanitation service delivery may be affected by the ability to commit to long-term processes from both development organizations (due to short funding cycles) and local leadership (due to political cycles).
- Sustainability-focused support activities – such as advocating for local government involvement, guiding the local government to allocate resources for sanitation over time, supporting reliable monitoring and evidence-based innovation, or agreeing on provision of continuous support with local partners – likely support the longevity of sanitation improvements.
- Reliable monitoring and targeted advocacy should be maintained over a long period to achieve structural change and leverage budgets, and ultimately provide adequate resource allocation for sanitation. Consequently, development organizations may consider implementation of sustainability activities after the local government becomes the sole facilitator for sanitation service delivery.

## 6. Conclusion

Our data suggests that sustainability factors identified through this study – including poverty, geography, social cohesion, population density, political will, and local government leadership – may have implications on where certain programmatic approaches will work, and where adaptations may be required. We found that successful systems strengthening with the commitment and buy-in of local governments, alongside more flexible, responsive, and long-term programming, may be required to achieve and maintain long-term access to sanitation. Further research and collaboration with development partners and local governments is needed to more fully understand the impact of these sustainability factors on rural sanitation program design.

## Data Availability

Data may be available if requested via email.

## Acknowledgments

This research is supported by SNV. The Sustainable Sanitation and Hygiene for All program is supported by the UK Department for International Development in Ethiopia, Uganda, Ghana, Zambia, Kenya, Mozambique, Tanzania, Nepal; the Australian Governments Department of Foreign Affairs and Trade (DFAT) in Nepal and Bhutan; the Stone Family Foundation in Cambodia; and the Embassy of the Kingdom of the Netherlands in Indonesia. We give special thanks to Antoinette Kome, Gabrielle Halcrow, Anne Mutta, Fanuel Nyaboro, Joseph Oluoch, Michael Bomji, Samson Wachara, Bendy Kipchoge, Kumbulani Ndlovu, Chainga Zulu, Edgar Chaamwe, Gian Melloni, Krishna Hari, Ugyen Rinzin, Kencho Wangdi, Raj Kumar, and to the data collection teams.

## Author Contributions

ZS and MCF conceptualized the study. ZS and EAU conducted the analysis and wrote the first draft. ZS, EAU, JSS, JVG, and MCF reviewed and edited the final draft. All authors read and approved the manuscript for publication.

## Ethics approval and consent to participate

The study authors were engaged by SNV as external evaluators to complete the study data. The Institutional Review Board of Emory University deemed the study exempt from review.

